# Closed-loop Neuromotor Training System Pairing Transcutaneous Vagus Nerve Stimulation with Video-based Real-time Movement Classification

**DOI:** 10.1101/2025.05.23.25327218

**Authors:** Minoru Shinohara, Arya Mohan, Nathaniel Green, Joshua N. Posen, Milka Trajkova, Woon-Hong Yeo, Hyeokhyen Kwon

## Abstract

As an emerging neurostimulation for improving motor rehabilitation, applying vagus nerve stimulation (VNS) after successful movement during training facilitates motor recovery in animals with neuromotor impairment. To translate this procedure to human rehabilitation in a non-invasive, objective, and automated manner, real-time classification of movement quality on a trial-by-trial basis in a minimally constrained state is required. In this work, we developed an integrated closed-loop system using video-based real-time movement classification that can automatically trigger transcutaneous VNS (tVNS) wirelessly as soon as successful movement is detected. We also created a film-like conformable tVNS electrode to be attached over the outer ear. For movement training, we focused on the use case of dance therapy (backward walking), which is widely used for people with Parkinson’s disease and older adults. Our markerless video analysis model could detect steps with 0.91 precision and 0.72 recall and classify successful backward steps with a 0.93 F1 score. The classification triggers tVNS through Bluetooth Low Energy communications with a trigger relay device we created. The integrated system enabled real-time automated classification and stimulation, triggering tVNS with 71.3% of the successful movements and taking 2.24 s from video capture to tVNS. We consider our work to be an important step toward patient-driven rehabilitation at home showcasing non-invasive, low-cost, and automated closed-loop neurostimulation technologies.

## 1 Introduction

The leading neurological causes of motor impairment in the adult population include stroke and Parkinson’s Disease (PD) [27, 36]. Individuals with motor impairments due to neurological disease or injury undergo rehabilitation training in the hope of recovering as much motor function as possible. The rate and extent of recovery with training varies from person to person due to various factors that influence neural plasticity [17, 24, 30]. Several types of neurostimulation interventions have been studied to facilitate neural plasticity and motor recovery, such as transcranial magnetic stimulation, transcranial direct current stimulation, and deep brain stimulation, which are applied before or during motor performance, without considering the trial-to-trial variability of movement quality during motor rehabilitation [34]. The current study developed a video-based system for an emerging neurostimulation intervention that requires trial-by-trial classification of movement quality.

Pairing brief vagus nerve stimulation (VNS) with motor training is an emerging intervention that has shown improved motor recovery in various neurological disorders, including human stroke survivors [9, 10, 20] and rats with stroke or spinal cord injury [11, 19, 25]. The scientific premise is the intriguing effect of stimulating the afferent vagus nerve, which transiently modulates neurotransmitters (e.g., norepinephrine, acetylcholine) in the cortex, which can facilitate or inhibit neuromotor adaptations, depending on the timing of stimulation in animal studies [6, 16]. Motor recovery or learning after multiple training sessions can be facilitated if VNS is applied only immediately after successful trials during motor practice. If the application of VNS is not associated with successful trials (e.g., random or delayed application), the facilitating effect cannot be obtained or turns into a negative effect [6, 11, 16]. It is assumed that transiently elevated neurotransmitters strengthen the synaptic efficacy of the recruited neural network. Therefore, it is critical to identify successful movements and apply VNS only after successful movements.

In translating this paradigm into human motor rehabilitation training, researchers surgically implanted a VNS electrode and device into the body of stroke survivors, and a therapist triggered the VNS application with a hand-held push-button by visually evaluating their movements and triggering VNS during specific movements in motor rehabilitation [9, 10]. While this form of intervention effectively facilitated motor recovery, it involves the risk and cost of the invasive procedure and the inconvenience and cost of the therapist classifying the movements. To overcome these problems, we aimed to develop a noninvasive and automated system for pairing VNS with motor training that does not require an invasive procedure or therapists’ manual inputs.

This development involved several aspects, both software and hardware. To enable automated movement classification for this purpose, we developed machine learning (ML) algorithms to identify and classify the quality of movement on a trial-by-trial basis in real time, using markerless motion capture with a standard low-cost video camera, such as a webcam. As an example of a movement to classify in this development of a closed-loop VNS system, we chose a backward stepping movement part of the adapted tango in older adults, which is used in dance therapy or training for individuals with stroke, PD, and even in healthy older adults [14, 38]. In the paradigm of applying VNS only after successful movements, avoiding misclassification of an “unsuccessful” movement as a “successful” movement (i.e., false positive) is critical and must be prioritized while misclassifying a “successful” movement as an “unsuccessful” movement (i.e., false negative) is tolerated.

**Figure 1:**
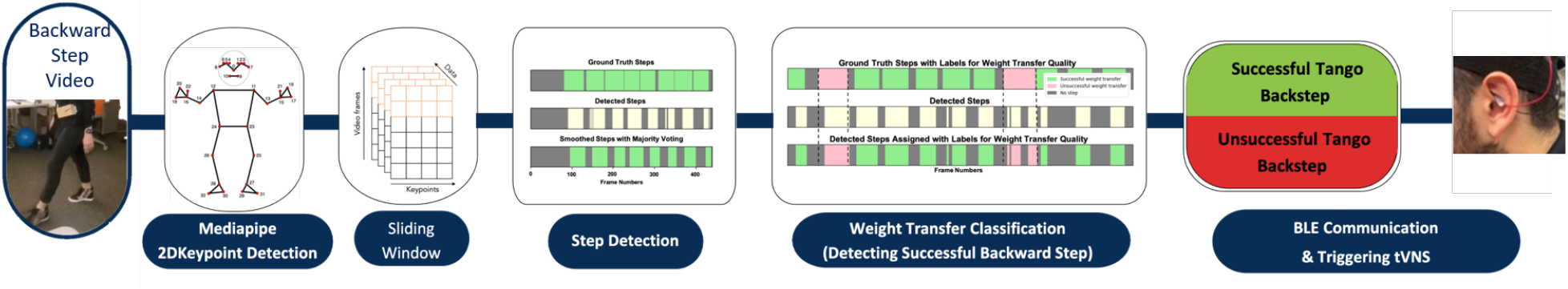
Overall pipeline combining video-based backward walking analysis and movement-associated transcutanous vagus nerve stimulation (tVNS). The machine learning (ML) pipeline detects backward steps from the frontal view videos and classifies the successful and unsuccessful backward steps. When successful steps are detected, tVNS is triggered for stimulation. Here, we show the view from the side view to better illustrate the backward step in our study design.

As an alternative to invasive VNS, we adopted a customized electrical stimulator for transcutaneous vagus nerve stimulation (tVNS) of the auricular nerve branch at the outer ear. Note that we have confirmed that tVNS can modulate the rate of motor learning in humans [32] as in invasive VNS. We also created a Bluetooth Low Energy (BLE)-based trigger relay device to wirelessly mediate video-based movement classification and tVNS application. To reduce the mass of the tVNS electrode on the outer ear, we developed a thin film-type conformable tVNS electrode that can reduce uncomfortableness and constraints resulting from wearing electrodes. Finally, we integrated the above-mentioned components into a real-time closed-loop system with automated movement classification pairing tVNS with successful movements, using a standard laptop computer with a built-in webcam. Although our original motivation is specific to a closed-loop tVNS system, the study results would be generally applicable to other use cases that require an automated VNS or tVNS system and real-time movement assessment with instantaneous feedback, which are key technologies needed for home-based rehabilitation.

## 2 Methods

### 2.1 Overall Scheme

The study collected markerless videos of older adults performing the tango backward walking using a standard camera. The frontal view of the videos was used to develop the ML algorithms. The video segmentation for each backward walking trial and labeling of successful and unsuccessful movements were manually annotated for developing ML systems.

A dataset was collected to develop ML-based step detection and classification algorithms to enable movement-associated tVNS during dance therapy. These algorithms and thin film-type tVNS electrodes were integrated into creating a closed-loop neuromotor training system for pairing tVNS with successful backward steps. The overall system pipeline is shown in Figure 1. The study was approved by the Institutional Review Board of the Georgia Institute of Technology (H23002).

**Figure 2:**
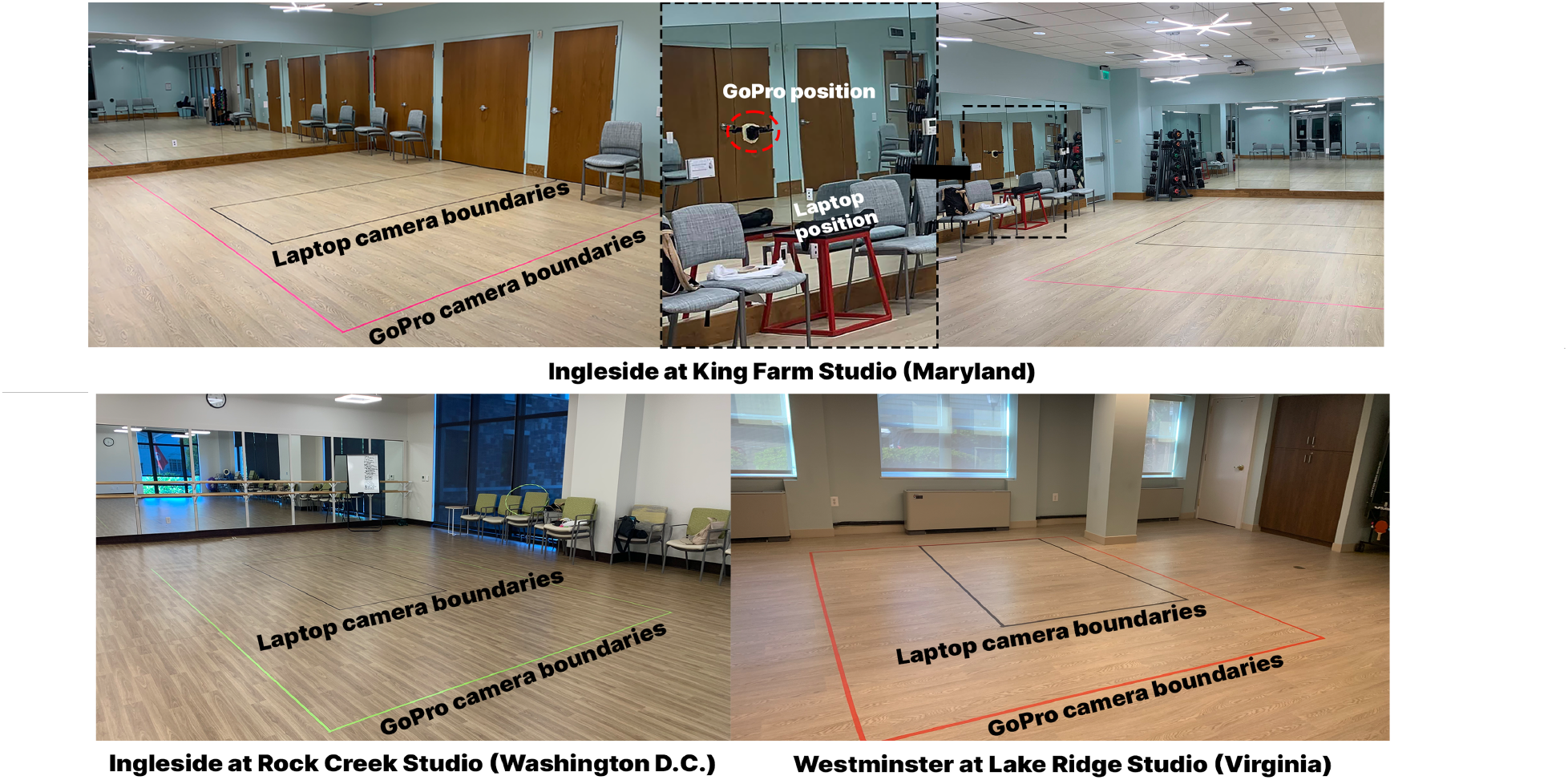
Camera and studio setup in the three locations of Ingleside Life-Plan Community

### 2.2 Human Data Collection

We collaborated with a life-plan retirement community, Ingleside, with three campuses in Washington D.C., Maryland, and Virginia in the USA, to collect human video data. Their executive team held orientation sessions to recruit and schedule participants.

#### 2.2.1 Participant Demographics

There were 28 unique participants, self-identified as female (N = 17) and male (N = 11) older adults with an age range of 73-93 years. Of these participants, 16 reported having a disability or impairment, such as motor/physical, visual, speech, cognitive, learning, neurological, auditory, or other. Among them, seven participants self-identified as having PD, with an average diagnosis duration of 6.1 (SD: 5.7) years. In 21 sessions across all three campuses, 56 repeating participants volunteered to engage. Data collection was completed within a month.

#### 2.2.2 Video Recording Set-Up

In all these communities, we collected markerless videos of their whole-body movements during adapted tango backward walking sessions with the participants. Figure 2 demonstrates the setup of the three different studios used in Maryland, Washington D.C., and Virginia. The studios were cleared of any objects in view, and the mirrors were covered to control for extraneous variables. We used a GoPro Hero 10 camera and also recorded sessions on a laptop as a backup device. The frame rates were set to 30 Hz for both devices. Participants were filmed utilizing three viewpoints: frontal, sagittal, and diagonal. In this work, we focused on using frontal-view movement analysis, which was considered to be easy to set up for subjects in home environments, compared to sagittal view. Frontal view is found to contain enough information to perform automatic recognition [3, 31].

#### 2.2.3 Movement Workshop Sessions

The data collection plan began with consulting a PD dance therapy expert to select the movements to be included. Research indicates that older individuals and those diagnosed with PD exhibit certain patterns of backward movement that suggest a reduction in mobility [13, 23]. A significant number of falls occur when moving in reverse. Therefore, it is crucial to allocate sufficient time and assistance for mastering the skill of backward walking, similar to the “tango walk.” In the Tango dance, while one partner advances forward, the other must navigate backward. This reverse movement in Tango is particularly beneficial for executing safe backward steps. Individuals should lean their bodies slightly forward towards their dancing partner and keep this forward-leaning stance consistently. Learners need to extend their toes backward first and then shift their hips carefully to align over their feet. Instructors guide their learners to avoid taking backward steps that cause their shoulders to lag behind their support base (beyond their toes). Whether moving forward or backward, the toe should be positioned beneath the hips and torso to ensure that the body’s center of mass is centered over the support base. The data collection procedures were as follows: 1) Participants were led through the informed consent process and were given an introduction to the project as well as made aware of all the data collection procedures, including video recording (5 min); 2) Participants engaged in a 45-min interaction session, which included a 5-min warm-up. They were introduced to the backward-walking tango movement by observing a movement demonstration three times, followed by a group practice; 3) Afterward, each participant performed the movement individually 10 times, and the performances were recorded as video data.

### 2.3 Video Preprocessing and Annotation

#### 2.3.1 Video Preprocessing

During the data preparation stage, the video data were documented and segmented into individual participant videos. These videos were identified by the campus ID, date and time of the session, movement type (backward walking, rhythm entrainment, or tango box), video view type (front, back, side, or diagonal), and participant ID. We manually annotated each video segment (start and end timestep) belonging to each backward step. Each backward step was also annotated with the movement being successful or not.

#### 2.3.2 Weight Transfer Quality Annotation

In the evaluation of whole-body motor skills, specifically in the context of assessing backward Tango walking movement, we utilized weight transfer, i.e., significant shifts in the center of mass as a participant shifts from one leg to the other, and foot alternation as criteria to determine the success of the observed movement. A review by Hasegawa [15] determined the most sensitive objective measures of balance dysfunction that differ between people with PD and healthy controls. Out of 93 measures, Anticipatory Postural Adjustments and dynamic posture were considered most sensitive for PD. Rather than manually calculating metric by metric, we chose a visual assessment approach by providing a set of criteria for video annotation. Visual assessments provide a holistic understanding of movement by capturing dynamic interactions between body parts, offering insights into the fluidity and smoothness of movements that isolated metrics cannot. ML tools excel in detecting subtle anomalies and complex patterns.

For a movement to be considered successful from a visual assessment, weight transfer must be clearly demonstrated by the participant shifting from one foot to the other without any shuffling, ensuring a distinct start and finish to the movement. This includes extending the back leg and bringing the feet together in a controlled manner. Similarly, for foot alternation to be considered successful, the participant must exhibit the ability to alternate feet for two full backward walking sequences. This involves stepping back with one foot, bringing the feet together, then stepping back with the opposite foot, and bringing the feet together again, ensuring a seamless transition between movements. The movement was labeled “successful” only when both criteria were satisfied. Conversely, the movement was labeled “unsuccessful” if either criterion was not met. The weight transfer criterion was not met when it involved shuffling or failed to clearly demonstrate initiation and conclusion, indicating imprecise weight transfer. A failure in foot alteration was marked by the inability to alternate between the right and left leg consistently, such as repeating the same leg for consecutive steps or incorporating a side-to-side backward motion, which deviates from the required technique of extending the leg straight back. These criteria served as fundamental measures in evaluating motor coordination and balance, contributing to a comprehensive understanding of the trial-to-trial variability of motor skill performance in each individual.

### 2.4 Video-based Step Detection and Weight Transfer Quality Classification from Frontal View

#### 2.4.1 Overview

Previous work demonstrated that ML-based approaches are effective for movement assessments[12]. In this work, we also applied ML to detect each step trial from a frontal view and classify successful and unsuccessful backward steps, which will later be communicated to tVNS over BLE, as demonstrated in Figure 1. Unlike previous work, we aimed to develop a lightweight ML pipeline that can process video data in real-time on low-cost commodity devices, like laptops, that have limited computing resources. To this end, we also applied feature selection techniques to downsize the models at each pipeline. This was to support our system to trigger tVNS immediately (or near real time) after a successful backward step and to be applicable for at-home rehabilitation.

#### 2.4.2 Pose Estimation and Preprocessing

The frontal view video was processed with Mediapipe [5] for estimating two-dimensional (2D) poses while subjects make multiple trials of backward steps. Mediapipe is specifically designed to provide estimation of real-time poses. The estimated poses provide 2D locations of 33 body keypoints additionally with the distance of each key point from the camera.

Extracting 2D poses from video in real-time preserves the entire pipeline’s privacy by not requiring the storage of raw video data. It also helps reduce the computational cost for the downstream step detection and weight transfer classification compared to directly processing raw video data. The extracted pose data are further preprocessed to normalize for subjects’ distances from the camera and varying heights of subjects. This is important as the subjects take multiple backward steps. They move further from the camera, making them smaller in the scene, which makes it challenging to identify subtle but important movements related to compensatory motions while making backward steps. Also, normalizing the subjects’ heights helps the downstream tasks to be invariant to the subjects’ heights.

**Figure 3:**
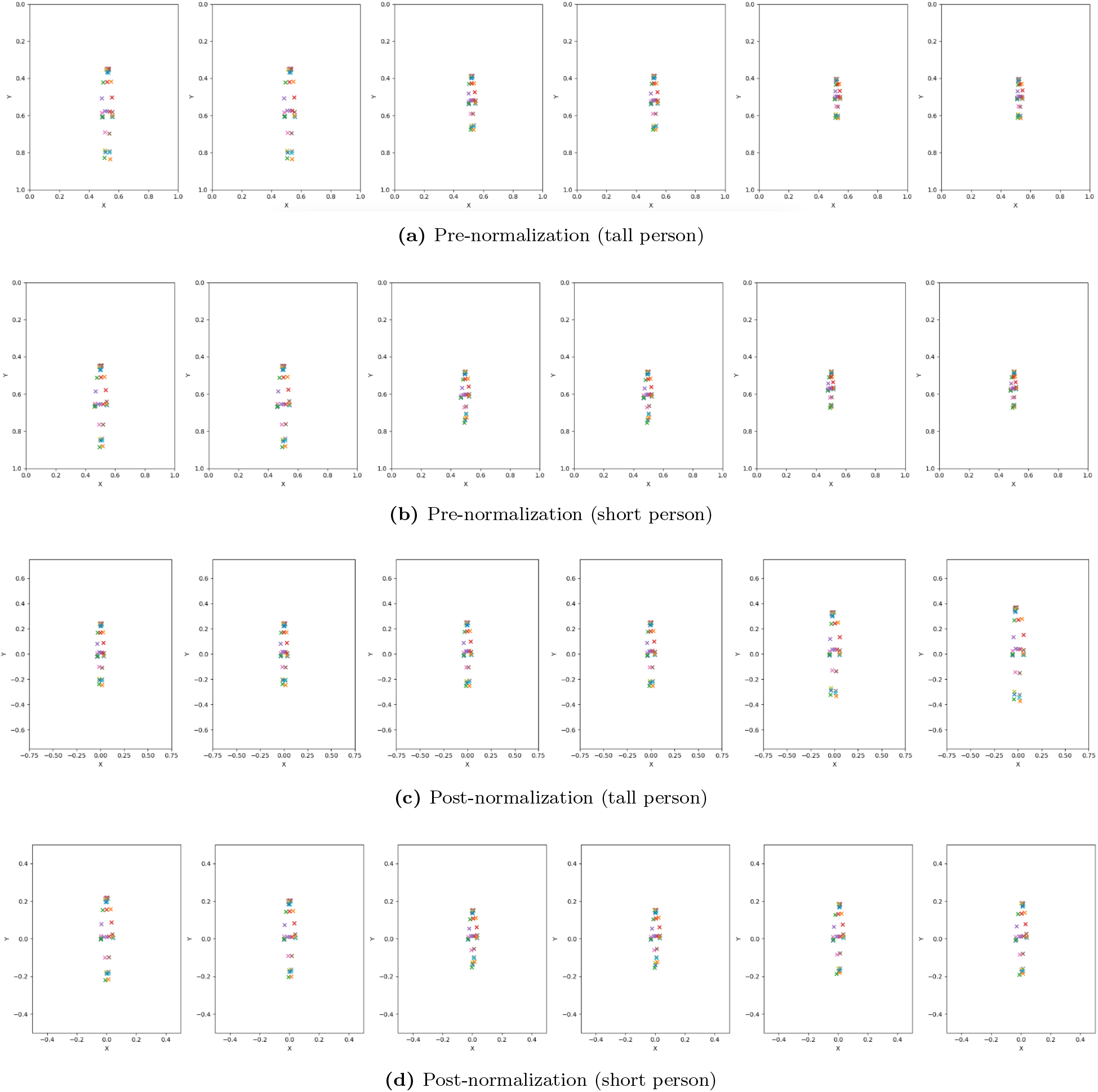
An example showing the normalization of pose keypoints of participants with different heights during backward steps. The panels illustrate the normalization process for a tall person before (a) and after (c) normalization, and a short person before (b) and after (d) normalization. This helps our downstream ML models learn the movement characteristics invariant to subject’s heights and distances from the camera.

Specifically, we normalized the size of 2D poses detected in each frame with the distance of the hip, which is the body center, detected by Mediapipe. This operation is similar to zooming in when the subjects are further away from the camera. The distance-normalized poses at each frame are further scaled with average height across all our subjects in the training dataset. This height-normalizing parameter was reused for test subjects. Figure 3 shows the before and after distance and height normalization applied for tall and short people in our subjects while taking backward steps. Before normalization, tall (Figure 3a) and short (Figure 3b) people show significantly varying pose sizes across subjects and frames within subjects. After normalization, both tall (Figure 3c) and short (Figure 3d) subject data showed consistent pose sizes across the frames. In our analysis, we found this normalization particularly important for the generalization of our downstream tasks across varying subjects.

#### 2.4.3 Step Detection

Conventionally, step detection tracks heel or toe markers to identify foot strikes for segmenting time windows for each step cycle, which is normally effective for sagittal views in the flat plane [18, 33]. However, these methods fall short when applied to frontal view analysis due to the ambiguity of exact foot contact with the ground, especially when a subject has a movement disorder and makes multiple compensatory movements and walks further from the camera. In Figure 4, the peak detection method for detecting the lowest heel marker for estimating heel strikes does not align sufficiently well with the ground truth. Multiple compensatory movements during backward steps produce multiple peaks in heel movements that are irrelevant to foot contacts.

To tackle the complex movements while making backward steps, we applied an ML-based approach to capture intricate movements and subtle nuances in step patterns from a frontal view that can generalize across heterogeneous subjects in this study having various kinds of motor impairment mentioned in Section 2.2.1. Following the standard practice in human activity recognition (HAR) pipeline [7], our step detection pipeline included sliding window, feature extraction, model training and inference, and post-processing. We chose a 30-s window size, which was the average step window size across all our subjects, with a 6-s interval.

If the last timestep of the window belongs to a ground-truth step window, we consider it part of a step segment. This was to ensure that our sliding window-based ML model could detect the onset and offset of a step segment as soon as it observes the relevant pattern in real time. From a window, we extracted the following set of gait features, including freezing index, central frequency, variance, and sample entropy, which were useful for characterizing gait patterns in older adults with PD [12, 21]. Although adapted tango backward steps are different from gait, we found those features are particularly useful for identifying characteristic movements in lower limbs in our older adults with heterogeneous conditions. Additionally, we included frequency and statistical time-series features widely used in HAR research [7], including aggregated autocorrelation, binned entropy, Fourier coefficients, and empirical cumulative distribution. The extracted features were used to train Random Forest and XGBoost classifiers, which were successfully used for HAR and gait analysis in previous work and also light-weighted for real-time processing on laptops [1, 22]. Lastly, the detected step segments are post-processed using majority voting-based smoothing with the window size equal to the median length of consecutive non-step segments. This was to remove or merge step segments that were too short detected by the proposed ML model, which is unrealistic. In our implementation, the sliding window-based step detection and majority voting-based smoothing are performed at the same time to ensure real-time analysis.

**Figure 4:**
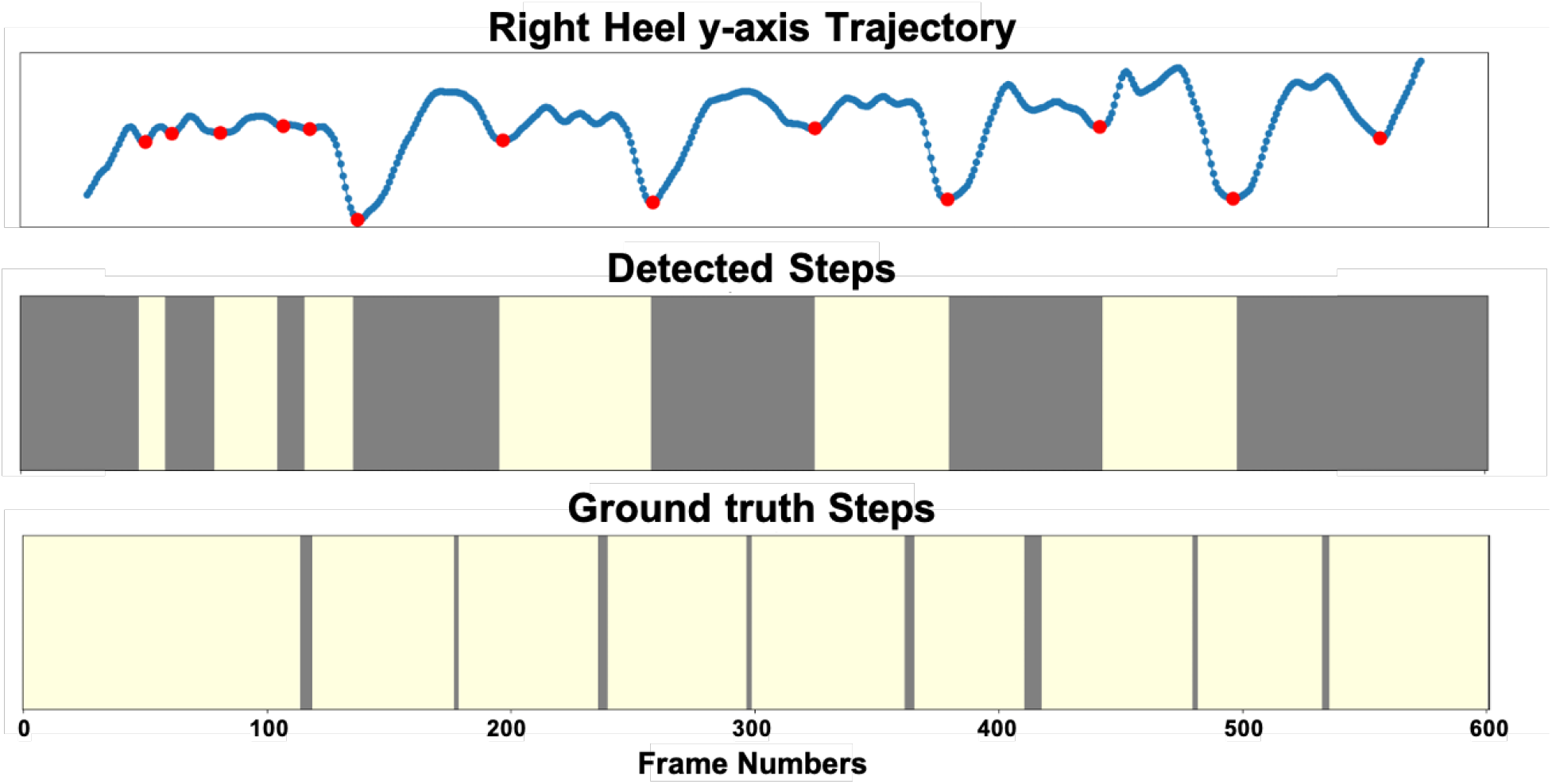
An example showing the comparison of traditional peak detection and actual step occurrences. The red dots represent detected peaks corresponding to heel strikes (top panel). The detected backward steps (yellow windows in the middle panel) do not align accurately with the true backward steps (yellow regions in the bottom panel).

#### 2.4.4 Backward Step Quality Classification

The detected step windows were further processed to classify between successful and unsuccessful backward steps. Here, we defined our task as to classify if the detect step window has any frames from unsuccessful backward steps. Since the step window detection likely does not perfectly align with the ground truth step window, some detected step windows can include both successful and unsuccessful backward steps. Therefore, the detected steps (Figure 5, middle) were labeled as either successful or unsuccessful backward step windows (Figure 5, bottom) based on the overlaps with ground truth step windows (Figure 5, top) with weight transfer quality annotation from Sec. 2.3. This was to ensure that our model could detect the step windows with purely successful backward steps, trigger tVNS, and maximize the rehabilitation effect. We take the keypoint time series from the detected step window for the classification task, from which we extract the same feature sets from the step detection task. The extracted features were input to Random Forest and Gradient Boosting models to classify the assigned successful or unsuccessful backward step labels.

**Figure 5:**
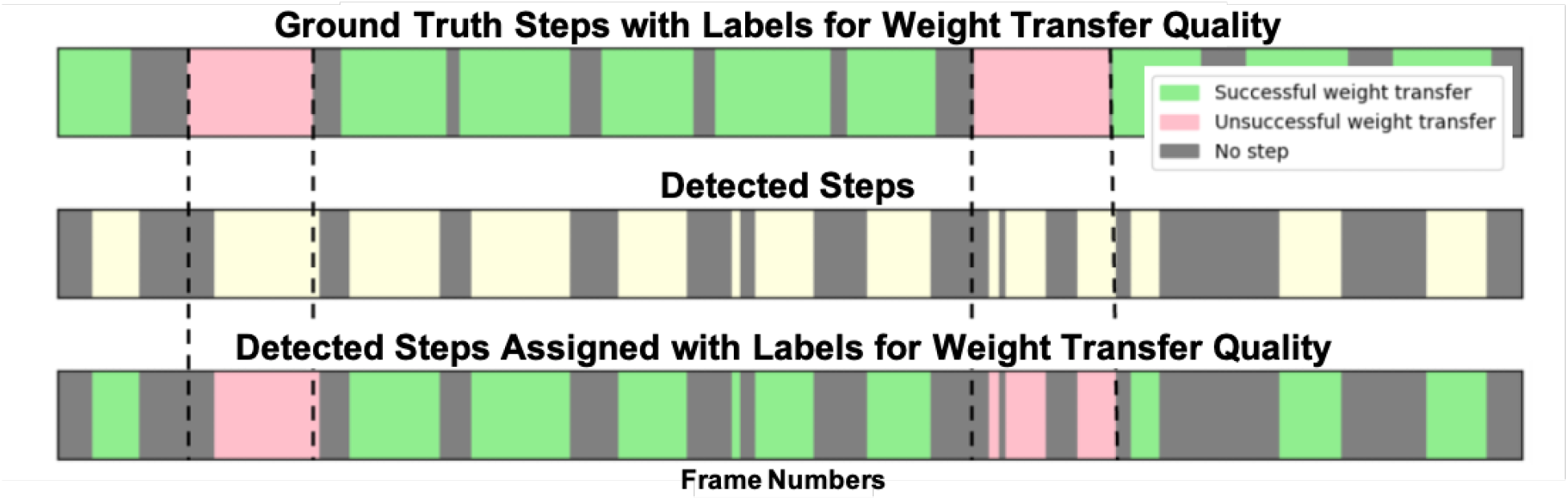
Assigning weight transfer quality labels for the detected steps based on the overlaps with ground truth step windows with the corresponding labels.

#### 2.4.5 Feature Selection

We employed permutation importance as our feature selection approach [2]. This method helps to identify and rank the features that contribute the most to the predictive power of our models. Permutation importance involves shuffling the values of each feature and measuring the impact on the model’s performance. The greater the decrease in performance, the more important the feature is.

Figure 6 illustrates the top 10 most important features identified using permutation importance. These features were crucial in our model’s ability to accurately detect steps and classify weight transfer quality. Notably, features like the central frequency of joints.right pinky.z and the maximum value of joints.neck.y emerged as significant contributors. In addition to the top features, we also analyzed the importance of all features through a heat map, as shown in Figure 7. This heatmap provides a comprehensive view of how different features from various keypoints contribute to the model’s performance. The color intensity indicates the mean decrease in performance when a feature is permuted, with darker colors representing higher importance.

Our feature selection process highlights the balance between maintaining model accuracy and achieving computational efficiency. By focusing on the most impactful features, we reduced the feature set size, leading to faster processing times and more efficient real-time analysis.

**Figure 6:**
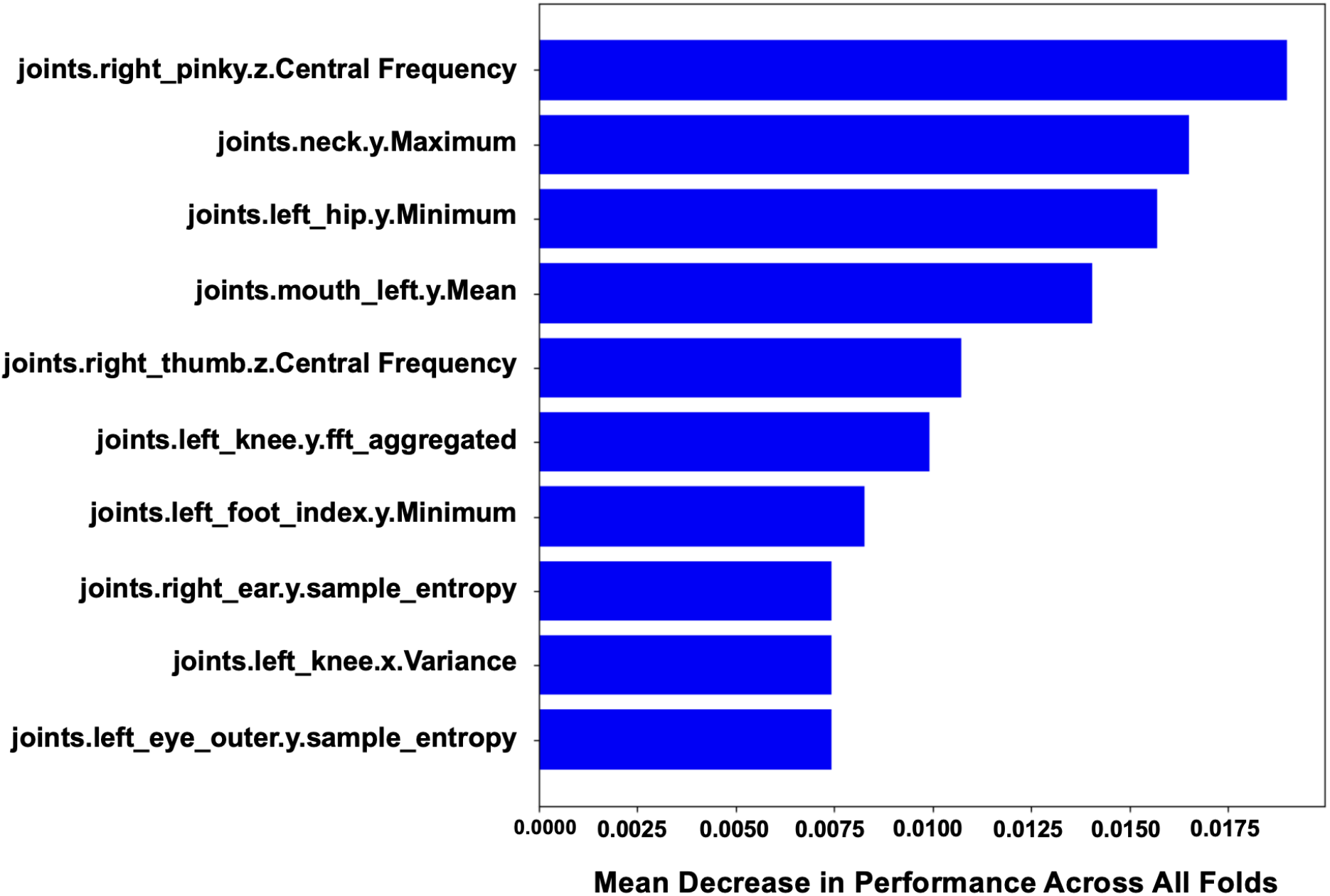
Top 10 most important features across all folds.

**Figure 7:**
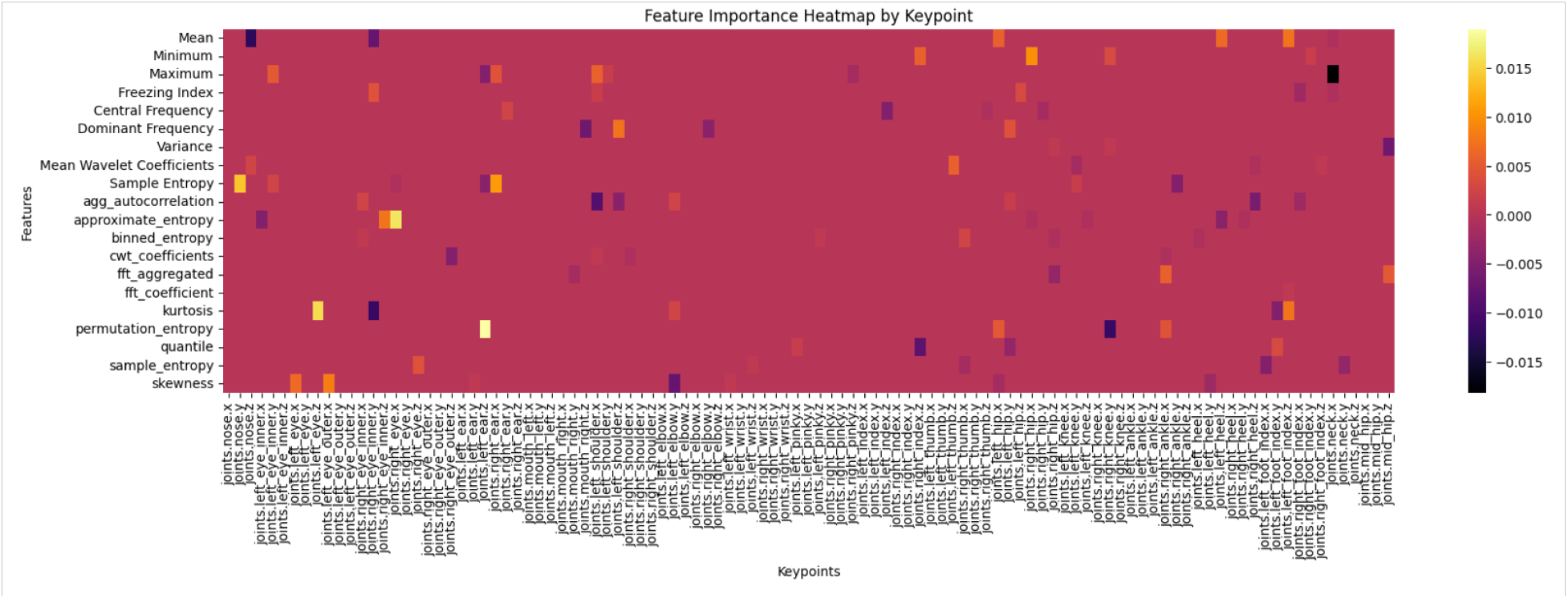
Feature importance heat map by keypoint.

#### 2.4.6 Evaluation

Both step detection and weight transfer quality classification were evaluated with user-independent 10-fold cross-validation, where the training, validation, and test sets have different user sets. We split our 60%, 20%, and 20% subjects into train, validation, and test sets, respectively. This was to evaluate the generalizability of our method when the trained model observes a new user’s movement. Both step detection and weight transfer quality classification models used the same split for train and evaluation to avoid information leaks. As mentioned earlier, we evaluated Random Forest and XGBoost models for both tasks. Although both tasks share an experiment protocol, the main evaluation metrics differ, which is explained below. In all our evaluations, we used 95% confidence intervals to statistically compare the significance of performance differences between models.

##### Step Detection

We first matched the ground truth and detected step segments based on the most overlapping time duration. Then, the paired ground truth and detected step segments were considered true positives. The unpaired ground truth and detected step segments were considered false negatives and false positives, respectively. Then, recall was used as the main metric for evaluating step-detection performance. Our goal was to identify as many true steps as possible, even at the expense of some false positives. In our deployment scenarios, missing a few steps (false negatives) was acceptable since subjects undergo multiple trials.

##### Weight Transfer Quality Classification

We prioritized Sensitivity (true positive rate). This metric is crucial because the tVNS system should activate only when a successful weight transfer is detected. Minimizing false positives is essential to maximize the rehabilitation effect. We also evaluated the ratio of true positive weight transfer classification from the detected steps based on ground truth successful backward steps, Successful Stimulation Rate 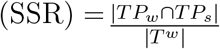, where |*TP*_*w*_ ∩ *TP*_*s*_| is the number of detected steps, *TP*_*s*_, with true positive weight transfer, *TP*_*w*_, and *T*_*w*_ is total number of ground truth steps with successful weight transfer. SSR directly translates to the successful stimulation rate of the proposed closed-loop video-based tVNS system.

In our experiment, we evaluated two scenarios, either using ground-truth step windows or detected step windows. Ground-truth step windows were used to evaluate our system in the ideal situation with perfect step detection. Specifically, we followed the implementation in our previous work [35]. We extracted 81 features from the keypoints in step windows, including all the angles and distances from the plumb line (or the center of gravity) - see Figure 8 and classified with 5-fold cross-validation using Gradient Boosting and Random Forest. When using the detected step window, we followed our approach mentioned earlier in Sec. 2.4.4.

##### Feature Selection

We also evaluated the performance of our models based on joint and features selected from our feature selection approach compared with when using full feature sets. We also compared the reduction of feature sets and inference time before and after feature selections. The computation time was measured on an Apple M1 chip with an 8-core CPU and an 8-core GPU times of inference on 30-s window size for step detection and weight transfer classification on multiple different sizes of detected steps.

**Figure 8:**
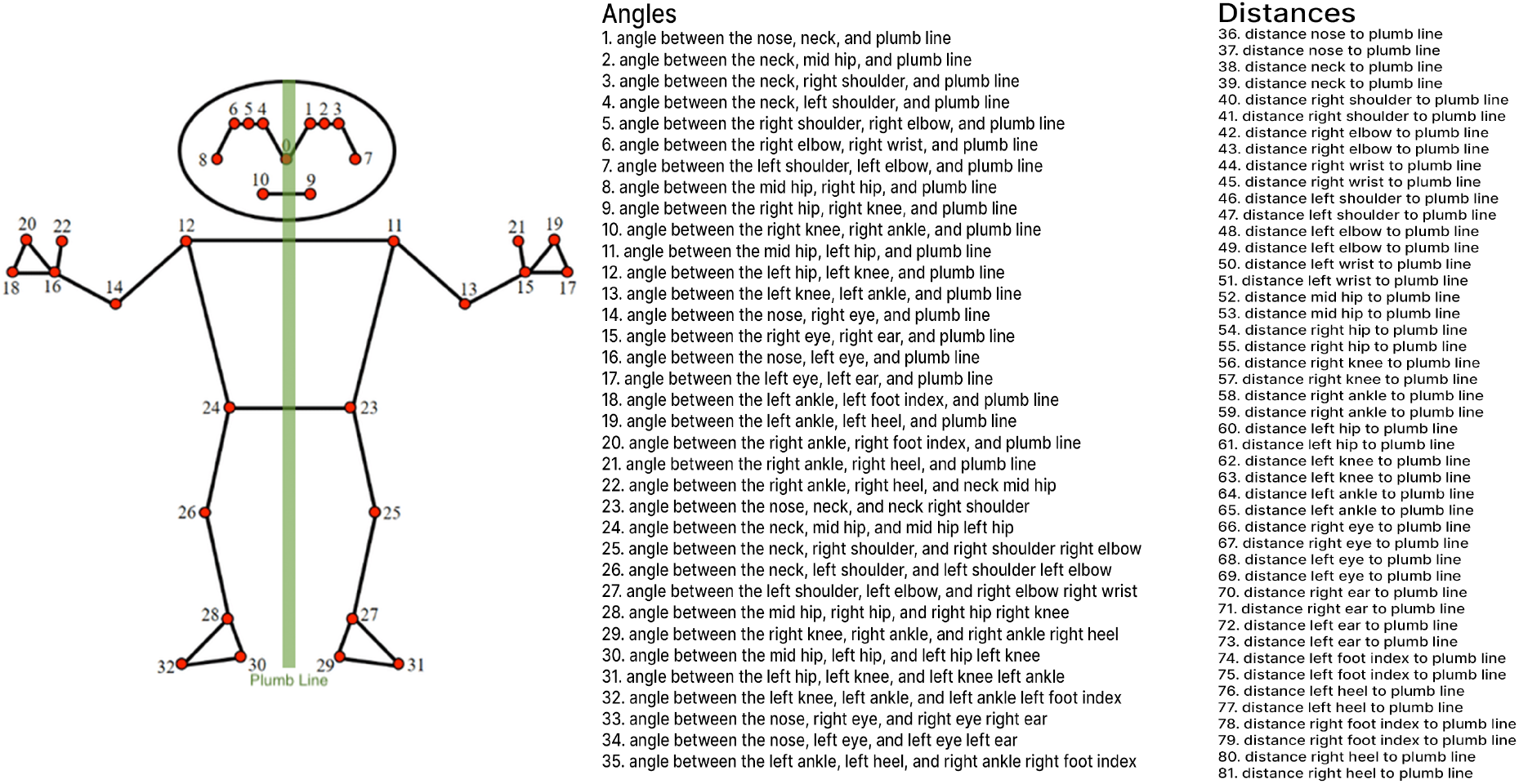
81 features including all the angles and distances from the MediaPipe key-body points.

### 2.5 tVNS Components

There were three aspects in our development of tVNS components. We 1) developed a film-type thin electrode that conforms to the shape of the tVNS target area in the outer ear (tragus or cymba concha), 2) adopted a commercially-available tVNS device customized to receive an external trigger signal, and 3) produced a trigger relay device between a computer and the tVNS device.

The film-type electrode was developed by producing a thin electrode with a serpentine pattern with gold [26]. It was created by first laminating a glass slide with a polyamide sheet. Subsequently, Au pellets were deposited onto the laminated slide, and a laser cutter was used to create a serpentine pattern modeled after the Peano design. The weight of the developed electrode was measured and compared with that of the commercially-available gel electrode (RELI-Stick, Soterixmedical, Woodbridge, NJ), which has dimensions of 9.5 x 12.7 mm and an outer adhesive of dimension 9.5 x 25.4 mm. Furthermore, the impedance of these electrodes was measured (1089NP Checktrode, UFI, Morro Bay, CA). To extend the lifespan of the film-type electrode, a long-lasting thin adhesive (Tegaderm 1622W, 3M, Saint Paul, MN) was applied. The gold pattern was then peeled off from the laminated film by the adhesive and stored until use. The adhesive was trimmed to fit the application area of the outer ear of the participant. A conductive path of Ag paint connected the electrode to a male pin connector, which was adhered to the back of the ear to offload the added weight of the wire and electrode. The connector allowed for a reusable wire to connect the stimulator to the attached electrode and disconnect at the end of testing.

We adopted a commercially-available tVNS stimulator (taVNS model 0125-LTE, Soterixmedical, Woodbridge, NJ) that was customized for our research purpose. With the customization, the stimulation can be triggered by receiving a 5 V square wave transistor-transistor logic (TTL) signal, and the stimulation parameters can be chosen and preset for a wider range. We also developed a trigger relay device, consisting of a microcontroller (Arduino Nano 33 BLE, Arduino, Somerville, MA) with a built-in BLE module and an LED. This device established a connection across the recording and classifying computer with a built-in webcam, the microcontroller, and the stimulator. The components of the trigger relay device are powered by a 6V rechargeable NiMH battery and housed in a 3D-printed box. We attached this trigger relay device and the stimulator by Velcro to a waist belt worn by the user. Upon receiving a predefined “success” command wirelessly via Bluetooth V4.0 BLE protocol, the device sends a 5 V digital signal to the stimulator via a detachable wire, which triggers tVNS applicaiton and turns on the LED to notify the stimulation application.

**Table 1:** Performance metrics for step detection models using different feature sets and models. The **Bold** text shows the best performances.

| Model | Features | Precision | Recall | F1 Score | True Positive Rate | False Positive Rate | Inference Time (ms) |
| --- | --- | --- | --- | --- | --- | --- | --- |
| Random Forest | Frequency | **0.92 ± 0.06** | 0.54 ± 0.17 | 0.66 ± 0.13 | 0.54 ± 0.17 | 0.30 ± 0.14 | 45 ± 9 |
| | Statistical | 0.88 ± 0.06 | 0.61 ± 0.07 | 0.72 ± 0.05 | 0.61 ± 0.07 | 0.27 ± 0.12 | 70 ± 8 |
| | Frequency + Statistical | 0.91 ± 0.04 | 0.65 ± 0.06 | 0.76 ± 0.03 | 0.65 ± 0.06 | 0.21 ± 0.08 | 82 ± 6 |
| | Frequency + Statistical (Top 10) | 0.81 ± 0.05 | 0.58 ± 0.07 | 0.63 ± 0.04 | 0.58 ± 0.07 | 0.28 ± 0.09 | 55 ± 5 |
| Gradient Boosting | Frequency | 0.89 ± 0.05 | **0.75 ± 0.06** | **0.81 ± 0.02** | **0.75 ± 0.06** | 0.16 ± 0.09 | 65 ± 7 |
| | Statistical | 0.84 ± 0.08 | 0.71 ± 0.11 | 0.76 ± 0.06 | 0.71 ± 0.11 | **0.46 ± 0.21** | 79 ± 12 |
| | Frequency + Statistical | 0.91 ± 0.06 | 0.72 ± 0.08 | 0.76 ± 0.13 | 0.74 ± 0.08 | 0.26 ± 0.05 | 85 ± 7 |
| | Frequency + Statistical (Top 10) | 0.88 ± 0.04 | 0.68 ± 0.06 | 0.69 ± 0.07 | 0.68 ± 0.06 | 0.29 ± 0.08 | 60 ± 8 |

**Table 2:** Performance metrics for weight transfer classification models using different feature sets and models. The **Bold** text shows the best performances.

| Model | Features | Precision | Recall | F1 Score | True Positive Rate | False Positive Rate | Inference Time (ms) | SSR (%) |
| --- | --- | --- | --- | --- | --- | --- | --- | --- |
| Random Forest | Frequency | 0.89 ± 0.10 | 0.58 ± 0.26 | 0.65 ± 0.23 | 0.58 ± 0.26 | 0.48 ± 0.30 | 63 ± 8 | 65.2 ± 4.5 |
| | Statistical | **0.91 ± 0.10** | 0.18 ± 0.17 | 0.26 ± 0.19 | 0.18 ± 0.17 | **0.16 ± 0.21** | 68 ± 7 | 61.5 ± 6.1 |
| | Frequency + Statistical | 0.84 ± 0.04 | 0.66 ± 0.06 | 0.73 ± 0.03 | 0.69 ± 0.06 | 0.24 ± 0.08 | 71 ± 6 | 71.0 ± 3.9 |
| | Frequency + Statistical (Top 10) | 0.72 ± 0.05 | 0.58 ± 0.08 | 0.67 ± 0.07 | 0.62 ± 0.08 | 0.26 ± 0.09 | 52 ± 4 | 64.9 ± 5.3 |
| Gradient Boosting | Frequency | 0.85 ± 0.09 | 0.72 ± 0.09 | 0.77 ± 0.07 | 0.72 ± 0.11 | 0.60 ± 0.18 | 68 ± 10 | 69.1 ± 5.0 |
| | Statistical | 0.81 ± 0.04 | 0.45 ± 0.23 | 0.54 ± 0.19 | 0.45 ± 0.23 | 0.52 ± 0.15 | 78 ± 11 | 63.7 ± 4.2 |
| | Frequency + Statistical | 0.88 ± 0.06 | **0.75 ± 0.08** | **0.81 ± 0.12** | **0.79 ± 0.04** | 0.17 ± 0.09 | 75 ± 6 | **71.3 ± 4.8** |
| | Frequency + Statistical (Top 10) | 0.76 ± 0.05 | 0.70 ± 0.07 | 0.76 ± 0.09 | 0.70 ± 0.07 | 0.19 ± 0.07 | 52 ± 7 | 66.2 ± 5.1 |

The tVNS parameters are adjustable on the stimulator and defined before task performance, including the frequency (30 Hz), pulse width (0.1 ms), and pulse train duration (500 ms) of the stimulation as constant values. The tVNS intensity could be set between 0.5 and 5 mA by 0.1 mA increments. Each individual has a unique intensity level defined by their pain threshold to tVNS. We define the task stimulation intensity as one decrement (0.1 mV) below the individual’s pain threshold.

### 2.6 System Integration and Implementation

#### 2.6.1 Integration

We integrated the software and hardware developed above to create and implement a closedloop system for pairing tVNS with successful movements during dance therapy. A simple web application user interface built in React was created to capture video data from the user’s default webcam via the browser. The user selects a trained model to load into the application for classification. Using Mediapipe, body keypoints are extracted from the incoming video stream and sent to a Python Flask server using the WebRTC protocol. The necessary input transformations are performed on the incoming stream of data and then sent to the chained step detection and weight transfer classification models wrapped as a single MLflow instance. The classification results are then sent back to the web UI via WebRTC, where the result is displayed to the user. Finally, the Web Bluetooth API (currently only supported in Chrome and Edge) is used to broadcast the classification result in the required format to a connected Bluetooth device. The web application and server are packaged with Docker for ease of deployment across various systems. Because user devices can vary widely in terms of their camera capabilities and processing speeds, the web UI takes a conservative approach to sending data to the server for classification to maximize portability across a range of potential devices and computing power. By default, the system throttles the incoming data stream to send data for classification every 0.25 s (4 Hz).

A standard laptop computer (MacBook Pro with a M1 Max processor, Cupertino, CA) and its built-in webcam were used to confirm the function of the integrated system. After a successful trial is observed by the classifying laptop, a string is sent to the paired Bluetooth module. The Bluetooth module is connected to a communication pin on the microcontroller. The communication pin is continuously read in search of the trigger command. When the trigger command (the ‘a’ character) is received, an output pin of the microcontroller is set to high for 1.0 s. The transition from a low (*<* 2 V) state to a high (5 V) state is received by the customized tVNS device (taVNS model 0125-LTE, Soterixmedical, Woodbridge, NJ) as a trigger. A pre-determined stimulation is then delivered to the participant.

#### 2.6.2 Evaluation

The real-time response delay of the integrated system was evaluated. The total delay of around 2 s was regarded acceptable to match the approximate effective delay to facilitate motor recovery according to an experimental study [11]. The total response delay (system delay) is composed of two elements, soft delay the time to acquire the incoming window of pose data and apply the step detection and weight transfer classification models and hard delay the time for the stimulation trigger to be generated and the subsequent stimulation felt by the user. In this evaluation, the initiation of system delay assessment was defined as the moment when a left backward step contacts the ground following a right backward step. A push button was placed and pressed with the left backward step to timestamp this moment for the evaluation purpose. The completion of the delay assessment was defined as the stimulation delivered to the user. This full evaluation of system delay was completed for 50 trials where the weight transfer model classified the steps as “good”. An Arduino with Bluetooth features was integrated with the acquisition software (Spike 2, Cambridge Electronic Design, Cambridge, UK) to record and determine the timing of analog signals at 10 kHz. The specific analog signals for quantification were the trigger signal for stimulation from the web app and the subsequent stimulation pulses at the electrodes relative to left foot contact. Note that the delays were only quantified when a button press was promptly followed by a stimulation trigger, indicating a “good” step was performed and the system classified the step accordingly.

**Figure 9:**
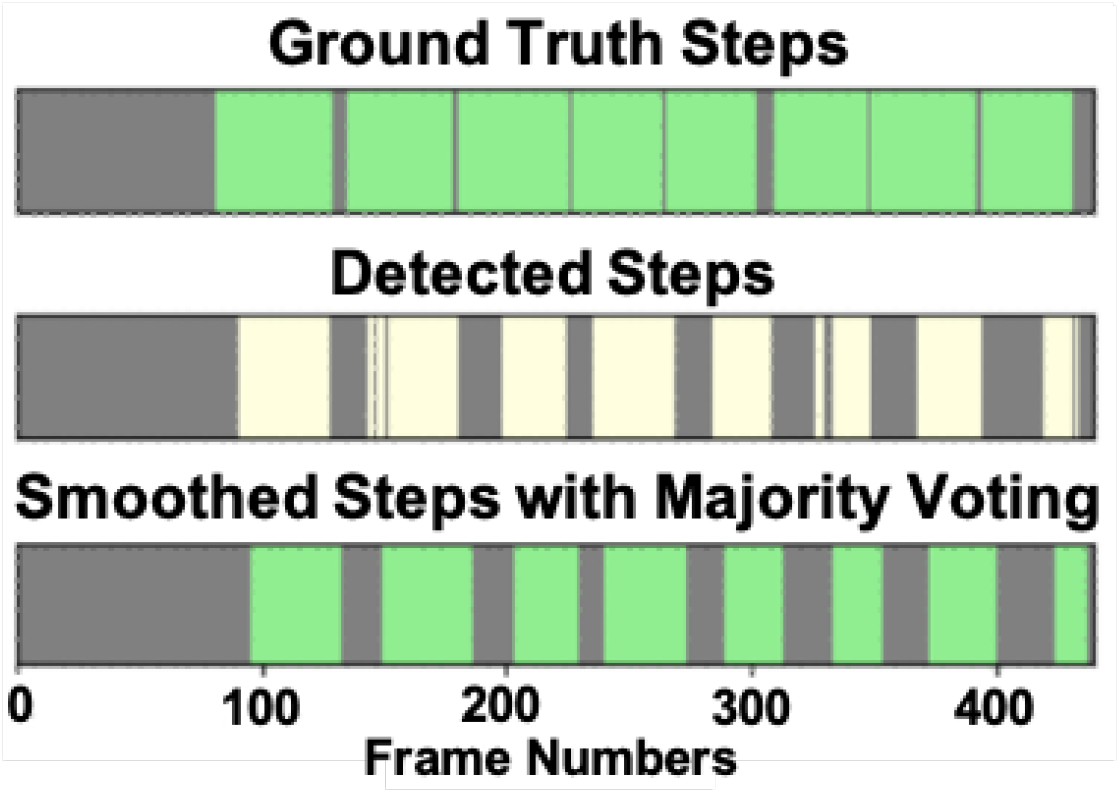
An example showing the ground truth steps (top), detected steps before (middle) and after majority voting (bottom)

## 3 Results

### 3.1 Characteristics of Source Data

The source data used for this study consist of the annotated video data described in Sec. 2.3, which were associated with the participant’s pose data derived using Mediapipe, comprising a total of 86 annotated video samples collected from the movement workshop sessions. Out of these, 523 backward Tango steps were successful trials, while 77 were unsuccessful.

### 3.2 Pose-Based Step Detection and Weight Transfer Classification from Frontal View

#### 3.2.1 Step Detection

As illustrated in Figure 4 (middle), traditional peak detection algorithms, which typically track the heel marker to segment steps, are shown to be insufficient for frontal view analysis with backward steps. In our experiment, the peak detection method showed an overall accuracy of 43.4%. Table 1 shows performances of the proposed ML-based backward step detection for different feature sets (frequency, statistical, and both) and models (Random Forest and Gradient Boosting). Overall, the Gradient Boosting model with frequency features showed the best performance, with a recall of 75%, which outperformed the best Random Forest model by absolute 10% recall. Figure 9 shows an example of where majority voting could smooth the detected step to merge or remove fragmented step windows that are very short.

#### 3.2.2 Weight Transfer Classification

When using ground truth step windows, Gradient Boost showed an accuracy of 87.9%, a precision of 90.2%, recall of 96.3%, and an F1 Score of 93.1%. Random forest produced similar results, showing an accuracy of 87.1%, a precision of 86.9%, a recall of 100.0%, and an F1 Score of 93.0%. This shows that in the ideal case when the step windows are perfectly detected, weight transfer quality can be reliably classified while taking backward steps. Table 2 shows the result for classifying weight transfer quality for our detected steps. The Gradient Boosting model using both frequency and statistical features was the best-performing model with the highest sensitivity (79%) and a low false positive rate (17%), showing that our model with noisy detected steps still contains sufficient motor signals in backward steps related to weight transfer quality. In this model, combining step detection and weight transfer showed the highest SSR (71.3%) with a higher true positive rate (79%) and a lower false positive rate in weight transfer (17%) compared with the Random Forest model using both frequency and statistical features, which showed a similar SSR (71.0%) but a lower true positive rate (69%) and a higher false positive rate in weight transfer (24%).

#### 3.2.3 Feature Selection

To ensure that our real-time analysis system is both efficient and effective, we employed permutation importance for feature selection. Following feature selection, we compared the performance metrics of our models before and after selecting the top features in Table 1 and Table 2. The performance comparison after feature selection as seen in Table 2 reveals that reducing the feature set to the top 10 most important features had a slight impact on the models’ performance with statistically similar performance considering confidence interval.

### 3.3 tVNS Components

The developed film-like conformable electrode had an active area dimension of 8.1 x 11.4 mm compared with 9.5 x 12.7 mm of the commercially-available gel electrode (Figure 10). It weighed 0.05 g at the active electrode portion, an 83% reduction in mass with only a 24% reduction in area, compared with 0.29 g for the commercially-available gel electrode. When an attached thin wire (7.9 cm) and a connector were included, the film-type electrode weighed 0.07 g compared with 0.43 g (84% reduction) for the gel electrode, including an attached wire of the same length. These are the main parts of the electrode attached to or around the outer ear of an individual. The film-type electrode had an impedance of 44.0 kΩ while the gel electrode had an impedance of 21.7 kΩ when attached to the skin surface. Since the stimulator has a cutoff impedance of less than 50 kΩ for “good” impedance, the film-type electrode was considered to have “good” skin contact with less mass compared with the commercially-available gel electrode. Perception and pain thresholds for each electrode were determined at the tragus of the left ear of an individual. The perception threshold was 0.6 mA and 0.8 mA for the film-type and gel electrodes, respectively. The pain threshold was 2.2 mA and 3.4 mA for the film-type and gel electrodes, respectively, while the maximum current of the stimulator is 5.0 mA.

The developed trigger relay device (Figure 11) included a microcontroller with integrated BLE communication, a LED indicator, a portable battery, and a container. The LED indicator was turned on when the device relayed a trigger signal from the computer to the stimulator. The device had dimensions of 8.9 x 8.9 x 2.9 cm (L x W x H) and weighed 213 g. The trigger relay device and the stimulator (15.9 x 7.3 x 1.6 cm, 137 g) can be attached on both sides of the hips by Velcro to a belt worn by a user for load balance. The output of the trigger relay device was connected to the tVNS device via a detachable wire connection.

**Figure 10:**
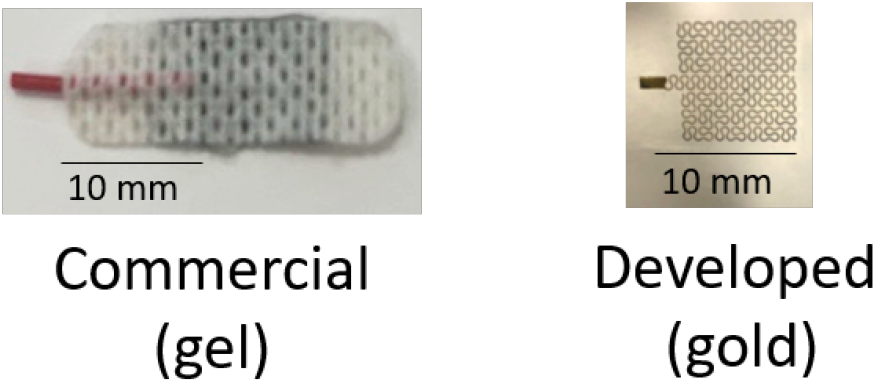
The commercially-available gel-type electrode (left) and the developed film-type gold electrode (right) for tVNS.

**Figure 11:**
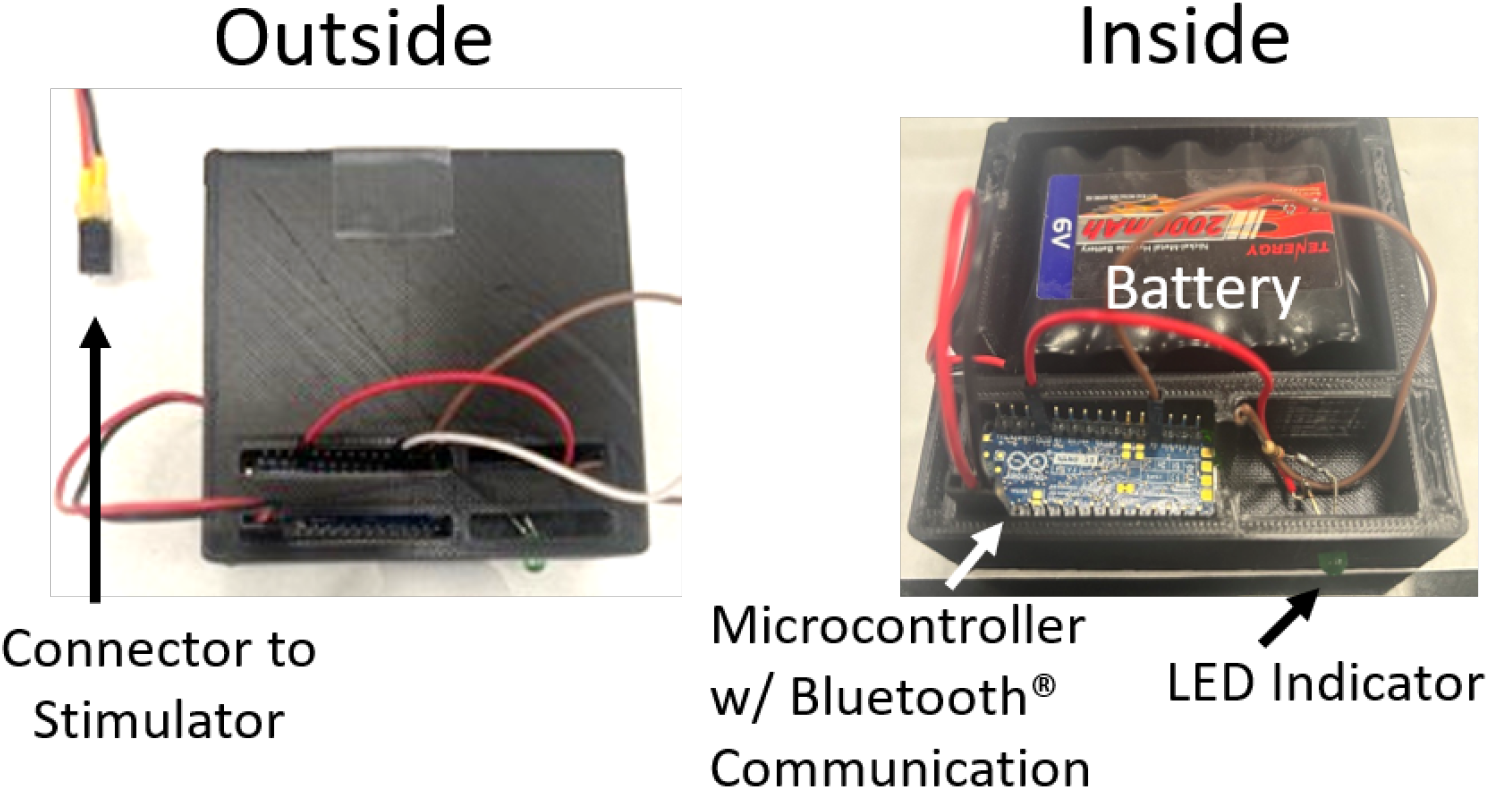
The developed trigger relay device (left: outside, right: inside)

### 3.4 Delays in Integrated Real-Time System

The hardware and software developed were configured into an integrated system (Figure 12). The delays in the entire real-time closed-loop tVNS system were evaluated from the completion of the backward step to the current stimulation applied to the user. As described above, the total delay and two sub-components (soft delay and hard delay) were measured to account for the delay from our model and the provided stimulation device, respectively. The total system delay was found to be 2.24 ± 0.37 s (mean ± SD), with a break down of 1.49 ± 0.37 s attributed to soft delay and 0.74 ± 0.03 s to hard delay.

**Figure 12:**
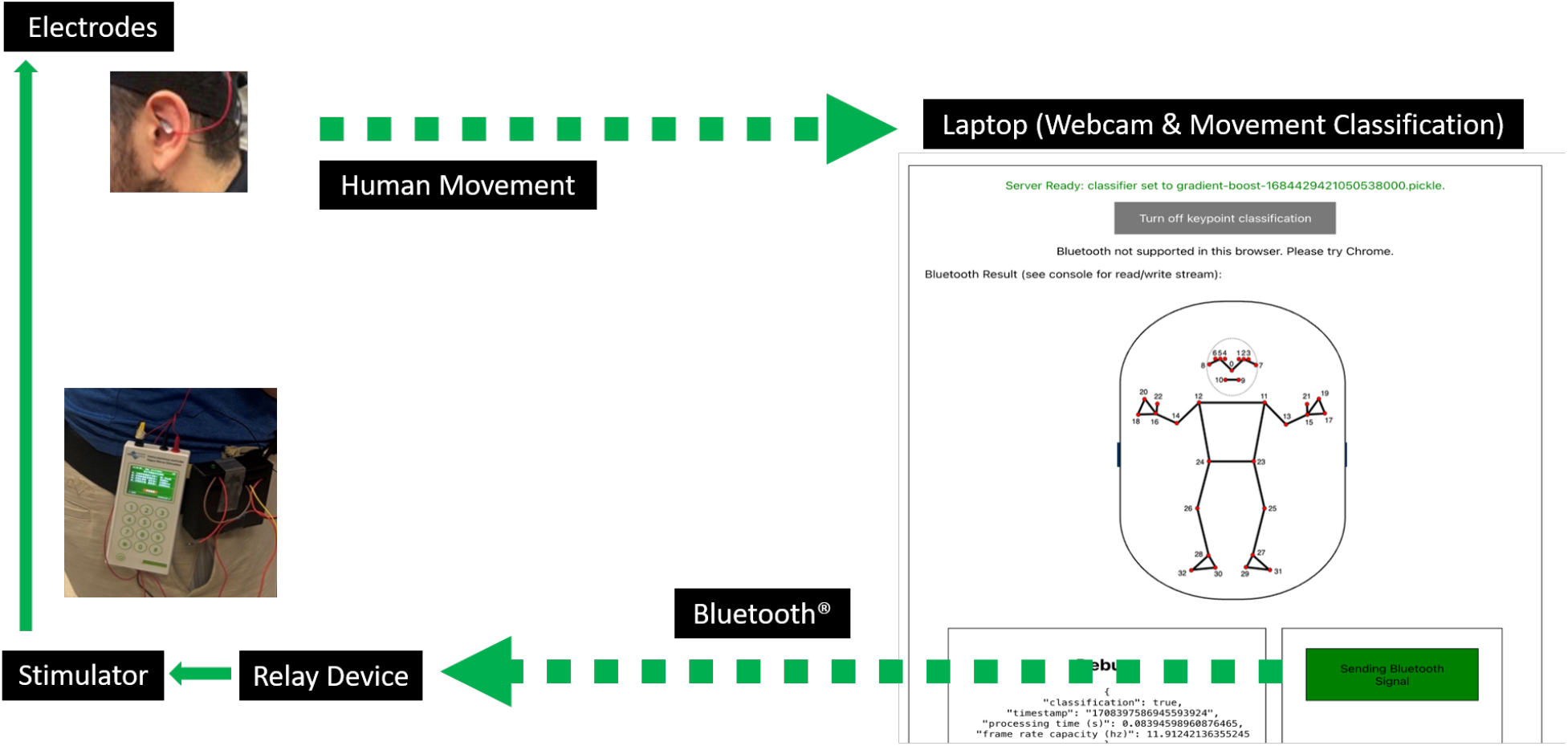
Configuration of the closed-loop neuromotor training system pairing tVNS (left) and movement classification (right). The arrows with a dotted line indicate wireless communication. Note that the trigger relay device (black box, left) is worn next to the stimulator in the photo for demonstration purposes, but it is intended to be worn on the other side for load balance.

## 4 Discussion

We developed a closed-loop neuromotor training system for pairing tVNS with successful movements using a standard camera. The major outcomes of the study are its development and validated high performance, which includes ML algorithms for detecting steps and identifying successful step trials in tango dance therapy using frontal-view video capturing in real time, a lighter film-type conformable tVNS electrode, and a trigger relay device that triggers tVNS application.

### 4.1 Step Detection and Weight Transfer Quality Integration

For step detection, the Gradient Boosting model outperforms the Random Forest model by 10%, demonstrating its capability to capture complex but subtle movement patterns while taking a tango backward step. Gradient Boosting can iteratively learn weak classifiers that capture high-level to low-level patterns to gradually reduce classification residuals. This was similar in weight transfer classification. The same Gradient Boosting model with combined features also performed best, achieving a recall of 75% and a low false positive rate of 17%. Overall system with leading to SSR of 71.3% underscores the model’s precision in applying tVNS only after successful weight transfers, which is crucial for effective therapeutic interventions. While the Random Forest model also showed a comparable SSR, its lower true positive rate and higher false positive rate in weight transfer are less favorable for applying tVNS at the appropriate timing.

Our observation showed that the most successful models could adapt to diverse movement styles and participant behaviors, indicating robustness across different scenarios. This was owing to Gradient-Boosting models’ iterative improvements in capturing diverse patterns across our participants. Models that performed poorly often struggled with rapid movements or atypical gait patterns, particularly in participants with PD, highlighting a potential area for improvement in future model training and development.

### 4.2 Film-Type Electrode and Trigger Relay Device

Historically, tVNS devices have been developed for their assumed application in a resting state to treat non-motor symptoms, such as anxiety, depression, and migraines [4, 29, 37]. The commercially available tVNS electrodes to be attached to the most appropriate part of the outer ear (i.e., tragus or cymba concha) include dry-type and gel-type electrodes, the former of which is unavailable in the USA. Since the mass of these electrodes at the small outer ear can cause uncomfortableness and constrain movements, we developed a lighter and conformable film-type tVNS electrode. We adopted our nanotechnology to create a thin electrode with a serpentine pattern with gold [26]. The achievement of less than 0.1 g mass with the film-type conformable electrode is a large improvement (greater than 80% reduction) compared with the gel electrode available in the USA. This light and conformable electrode can reduce the uncomfortableness of wearing something on the outer ear, especially during motor training. The impedance of the film-type electrode was higher compared with the gel electrode, but it is still below the cutoff impedance range (below 50 kΩ) in a commercially-available tVNS device (taVNS model 0125-LTE, Soterixmedical, Woodbridge, NJ). The intensity of tVNS is typically determined based on the perception or pain threshold in each individual [28, 32], while the maximum intensity is often limited for safety reasons (e.g., 5 mA for the taVNS model 0125-LTE). The lower thresholds with the film-type compared with the gel-type electrode allow more individuals to reach perception or pain thresholds within the limit.

The trigger relay device was created to wirelessly receive the timing of ML-based identification of successful movements and immediately trigger tVNS application. Since its dimensions and mass are similar to those of the tVNS device, wearing each device on each side of the waist will make comfortable, load-balanced attachments. The communication from the trigger relay device to the tVNS device is via a detachable wire connection due to the technical limitation of the commercially-available tVNS device. If a tVNS device with a Bluetooth capability becomes available, the trigger relay can be fully wireless with minor adjustments of the microcontroller program. The LED-based notification of tVNS timing with the device will allow an observer to monitor the execution.

### 4.3 Real-Time System Deployment

One obstacle to real-time deployment of such a system is processing the incoming data stream fast enough to produce a real-time result without creating a bottleneck. In the web user interface, Mediapipe is capable of extracting keypoints from incoming video rapidly, and the transfer of this data to the server hosting the classifier via WebRTC is also fast. However, performing the necessary pre-processing steps on this data stream and obtaining a classification result can add time to this process, affecting the ability of the system to respond in real time. Therefore, we needed to limit the amount of data sent to the classifier to avoid slowing down the entire pipeline. Depending on the needs of a particular classifier, it is certainly possible to optimize the data pipeline and tune it for a specific set of requirements. In a study examining the effect of VNS timing on its efficacy in facilitating motor recovery in rats with spinal cord injury revealed that substantial efficacy was evident when VNS was applied within approximately 2 s after each successful movement [11]. In the developed system, the total system delay was 2.24 s, which is around the approximated effective delay. In the current delay, 0.74 s was attributed to the device-dependent hardware delay within the commercially available tVNS device (taVNS model 0125-LTE). According to the manufacturer (Soterixmedical), this device-dependent delay can be reduced to less than 0.05 s with further customization to make the total delay less than 2 s if needed. Since the current system satisfied the approximate effective delay and also due to resource constraints and the required customization time for the device used in other ongoing projects, we decided to leave this option for a future opportunity. For the purposes of our prototype application, limiting the data stream to send updates every 0.25 s is more than sufficient to allow the system to deliver real-time classification results within this range. In practice, this updating speed could be increased depending on the user’s hardware. In testing on a modern laptop computer (e.g., MacBook Pro with M1 chip), processing speeds between 10-15 Hz are achieved and could likely be optimized much further depending on potential specific use cases.

### 4.4 Potential Applications

We developed this closed-loop system to automatically apply tVNS at the appropriate timing during motor training using a backward step example of dance therapy for older adults with and without PD. The developed system and its components can be applied to various use cases, including objective real-time movement assessment, automated result feedback, automated invasive VNS or noninvasive tVNS systems, home-based rehabilitation, and various disease and movement models. The movement classification and VNS triggering by a therapist can be replaced with our developed system. The classification results of the system will serve as a real-time objective movement assessment. The results of this ML-based classification can be utilized to automatically provide immediate feedback to an exerciser, such as auditory or visual cues. Additionally, the results can also be tallied across trials to automatically generate a score (e.g., success rate) for the training. In studies pairing invasive VNS with motor rehabilitation in post-stroke individuals, VNS was wirelessly triggered via Bluetooth by an on-site therapist visually classifying movements and pressing a hand-held trigger button [9, 10]. An automated invasive VNS or noninvasive tVNS system, which requires no therapist but just a laptop computer, will enable home-based rehabilitation. The type of movement to be trained and the associated classification criteria depend on the purpose and the condition (e.g., stroke, spinal cord injury) of the exerciser. The applicability of the developed system to various conditions depends on the availability of ML algorithms for classifying quality in targeted movements. The current ML algorithms are expected to be applicable, with minor adjustments, to other types of gait assessment. Also, with the latest advances in model compression methods for deep learning models [8], our system allows users to plug-and-play and replace ML models with more advanced models, significantly improving the detection performance for successful weight transfer. Our future studies will explore ML algorithms for other types of movement, such as upper-limb movement in individuals who have had a stroke.

## 5 Conclusion

This study developed and deployed a video-based closed-loop system for pairing tVNS with specific movements during motor training. This process included the development of ML algorithms for real-time classification of movement quality in dance therapy for older adults with and without PD, improvement from the gel-type tVNS electrode to a lighter film-like conformable electrode, creation of a trigger relay device, and system integration and implementation. It was demonstrated that the developed system in a mobile device with a built-in webcam is capable of identifying and classifying the quality of backward walking in dance therapy at high accuracy and automatically applying tVNS with a short delay (within 2.24 s) when appropriate.

## Data Availability

All data produced in the present study will be available upon reasonable request to the authors after peer-reviewed publication.

## 6 Acknowledgments

We would like to acknowledge the executive team and participants of the three campuses of Ingleside retirement community, located in Washington, D.C., Maryland, and Virginia in the USA. Additionally, we extend our gratitude to Madeleine Hackney for her invaluable assistance in identifying the movements to be studied in dance therapy. We also thank Koki Asahina, Naiji Gong, and Chanyeong Choi for their technical assistance.

This work was supported in part by the McCamish Parkinson’s Disease Innovation Program at Georgia Tech and Emory University. Hyeokhyen Kwon was partially funded by the National Institute on Deafness and Other Communication Disorders (grant # 1R21DC021029-01A1), NC NM4R 2024-2025 Pilot Project Grant (AWD-006196-G1), and the James M. Cox Foundation and Cox Enterprises, Inc., in support of Emory’s Brain Health Center and Georgia Institute of Technology.

## 7 Note

This article has been submitted to *ACM Transactions on Computing for Healthcare* for consideration. Its contents may not be further disseminated until a final decision regarding publication has been made and further permission has been granted.

## References

[1] Lidia Alecci, Leonardo Alchieri, Nouran Abdalazim, Pietro Barbiero, Silvia Santini, and Martin Gjoreski. Enhancing xgboost with heuristic smoothing for transportation mode and activity recognition. In Adjunct Proceedings of the 2023 ACM International Joint Conference on Pervasive and Ubiquitous Computing & the 2023 ACM International Symposium on Wearable Computing, pages 540–545, New York, NY, USA, 2023. Association for Computing Machinery.

[2] André Altmann Laura Toloşi, Oliver Sander, and Thomas Lengauer. Permutation importance: a corrected feature importance measure. Bioinformatics, 26(10):1340– 1347, 2010.

[3] Olivier Barnich and Marc Van Droogenbroeck. Frontal-view gait recognition by intra- and inter-frame rectangle size distribution. Pattern Recognition Letters, 30(10):893–901, 2009.

[4] Sebastian Bauer, Hartmut Baier, Christoph Baumgartner, Katrin Bohlmann, Susanne Fauser, Wolfgang Graf, Barbara Hillenbrand, Martin Hirsch, Christina Last, Holger Lerche, Thomas Mayer, Andreas Schulze-Bonhage, Bernhard Jochen Steinhoff, Yvonne Weber, A Hartlep, Felix Rosenow, and Hajo M Hamer. Transcutaneous vagus nerve stimulation (tvns) for treatment of drug-resistant epilepsy: A randomized, double-blind clinical trial (cmpse02). Brain Stimulation, 9(3):356–363, 2016.

[5] Valentin Bazarevsky, Ivan Grishchenko, Karthik Raveendran, Tyler Zhu, Fan Zhang, and Matthias Grundmann. Blazepose: On-device real-time body pose tracking. arXiv preprint arXiv:2006.10204, 2020.

[6] Spencer Bowles, Jordan Hickman, Xiaoyu Peng, W Ryan Williamson, Rongchen Huang, Kayden Washington, Dane Donegan, and Cristin G Welle. Vagus nerve stimulation drives selective circuit modulation through cholinergic reinforcement. Neuron, 110(17):2867–2885, 2022.

[7] Andreas Bulling, Ulf Blanke, and Bernt Schiele. A tutorial on human activity recognition using body-worn inertial sensors. ACM Computing Surveys (CSUR), 46(3):1–33, 2014.

[8] Pierre Vilar Dantas, Waldir Sabino da Silva Jr, Lucas Carvalho Cordeiro, and Celso Barbosa Carvalho. A comprehensive review of model compression techniques in machine learning. Applied Intelligence, 54(22):11804–11844, 2024.

[9] Michael J Darrow, Miranda Torres, Maria J Sosa, Tanya T Danaphongse, Zainab Haider, Robert L Rennaker, Michael P Kilgard, and Seth A Hays. Vagus nerve stimulation paired with rehabilitative training enhances motor recovery after bilateral spinal cord injury to cervical forelimb motor pools. Neurorehabilitation and Neural Repair, 34(3):200–209, 2020.

[10] Jesse Dawson, Charles Y Liu, Gerard E Francisco, Steven C Cramer, Steven L Wolf, Anand Dixit, Jen Alexander, Rushna Ali, Benjamin L Brown, Wuwei Feng, et al. Vagus nerve stimulation paired with rehabilitation for upper limb motor function after ischaemic stroke (vns-rehab): a randomised, blinded, pivotal, device trial. The Lancet, 397(10284):1545–1553, 2021.

[11] Patrick D Ganzer, Michael J Darrow, Eric C Meyers, Bleyda R Solorzano, Andrea D Ruiz, Nicole M Robertson, KatherineS Adcock, Justin T James, Han S Jeong, April M Becker, Mark P Goldberg, David T Pruitt, Seth A Hays, Michael P Kilgard, and Robert L 2nd Rennaker. Closed-loop neuromodulation restores network connectivity and motor control after spinal cord injury. eLife, 7:e32058, 2018.

[12] N Jabin Gong, Gari D Clifford, Christine D Esper, Stewart A Factor, J Lucas McKay, and Hyeokhyen Kwon. Classifying tremor dominant and postural instability and gait difficulty subtypes of parkinson’s disease from full-body kinematics. Sensors, 23(19):8330, 2023.

[13] Madeleine E Hackney and Gammon M Earhart. Backward walking in parkinson’s disease. Movement Disorders, 24(2):218–223, 2009.

[14] Madeleine E Hackney and Kathleen McKee. Community-based adapted tango dancing for individuals with parkinson’s disease and older adults. Journal of Visualized Experiments, (94):e52066, 2014.

[15] Naoya Hasegawa, Vrutangkumar V Shah, Patricia Carlson-Kuhta, John G Nutt, Fay B Horak, and Martina Mancini. How to select balance measures sensitive to parkinson’s disease from body-worn inertial sensors—separating the trees from the forest. Sensors, 19(15):3320, 2019.

[16] Seth A Hays, Navid Khodaparast, Andrea Ruiz, Andrew M Sloan, Daniel R Hulsey, Robert L Rennaker, Michael P Kilgard, et al. The timing and amount of vagus nerve stimulation during rehabilitative training affect poststroke recovery of forelimb strength. Neuroreport, 25(9):676–682, 2014.

[17] Brenton Hordacre, Mitchell R Goldsworthy, Ann-Maree Vallence, Sam Sarvishi, Bahar Moezzi, Masahi Hamada, John Rothwell, and Michael Ridding. Variability in neural excitability and plasticity induction in the human cortex: A brain stimulation study. Brain Stimulation, 10:588–595, 2017.

[18] Chrysostomos Karakasis and Panagiotis Artemiadis. Real-time kinematic-based detection of foot-strike during walking. Journal of Biomechanics, 129:110849, 2021.

[19] Navid Khodaparast, Seth A Hays, Andrew M Sloan, Daniel R Hulsey, Andrea Ruiz, Maritza Pantoja, Robert L Rennaker II, and Michael P Kilgard. Vagus nerve stimulation during rehabilitative training improves forelimb strength following ischemic stroke. Neurobiology of Disease, 60:80–88, 2013.

[20] Teresa J Kimberley, David Pierce, Cecília N Prudente, Gerard E Francisco, Nuray Yozbatiran, Patricia Smith, Brent Tarver, Navzer D Engineer, David Alexander Dickie, Danielle K Kline, et al. Vagus nerve stimulation paired with upper limb rehabilitation after chronic stroke: a blinded randomized pilot study. Stroke, 49(11):2789–2792, 2018.

[21] Hyeokhyen Kwon, Gari D Clifford, Imari Genias, Doug Bernhard, Christine D Esper, Stewart A Factor, and J Lucas McKay. An explainable spatial-temporal graphical convolutional network to score freezing of gait in parkinsonian patients. Sensors, 23(4):1766, 2023.

[22] Hyeokhyen Kwon, Catherine Tong, Harish Haresamudram, Yan Gao, Gregory D Abowd, Nicholas D Lane, and Thomas Ploetz. Imutube: Automatic extraction of virtual on-body accelerometry from video for human activity recognition. Proceedings of the ACM on Interactive, Mobile, Wearable and Ubiquitous Technologies, 4(3):1–29, 2020.

[23] Yocheved Laufer. Effect of age on characteristics of forward and backward gait at preferred and accelerated walking speed. The Journals of Gerontology Series A: Biological Sciences and Medical Sciences, 60(5):627–632, 2005.

[24] Virginia López-Alonso, Binith Cheeran, Dan Río-Rodríguez, and Miguel Fernándezdel Olmo. Inter-individual variability in response to non-invasive brain stimulation paradigms. Brain Stimulation, 7:372–380, 2014.

[25] Eric C Meyers, Bleyda R Solorzano, Justin James, Patrick D Ganzer, Elaine S Lai, Robert L Rennaker, Michael P Kilgard, and Seth A Hays. Vagus nerve stimulation enhances stable plasticity and generalization of stroke recovery. Stroke, 49(3):710–717, 2018.

[26] James J Norton, Dong Sup Lee, Jung Woo Lee, Woosik Lee, Ohjin Kwon, Phillip Won, Sung-Young Jung, Huanyu Cheng, Jae-Woong Jeong, Abdullah Akce, Stephen Umunna, Ilyoun Na, Yong Ho Kwon, Xiao-Qi Wang, Zhuanglian Liu, Ungyu Paik, Yonggang Huang, Timothy Bretl, Woon-Hong Yeo, and John A Rogers. Soft, curved electrode systems capable of integration on the auricle as a persistent brain-computer interface. Proceedings of the National Academy of Sciences, 112(13):3920–3925, 2015.

[27] Francisco Purroy and N Montalá. Epidemiology of stroke in the last decade: a systematic review. Revista de Neurología, 73(9):321–336, 2021.

[28] Jessica N Redgrave, Lucy Moore, Tosin Oyekunle, Maryam Ebrahim, Konstantinos Falidas, Nicola Snowdon, Ali Ali, and Arshad Majid. Transcutaneous auricular vagus nerve stimulation with concurrent upper limb repetitive task practice for poststroke motor recovery: A pilot study. Journal of Stroke and Cerebrovascular Diseases, 27(7):1898–2005, 2018.

[29] Peijing Rong, Jun Liu, Liping Wang, Rupeng Liu, Jilang Fang, Jingjun Zhao, Yufeng Zhao, Honghong Wang, Mark Vange, Sharon Sun, Hui Ben, Joel Park, Shaoyuan Li, Hong Meng, Bing Zhu, and Jian Kong. Effect of transcutaneous auricular vagus nerve stimulation on major depressive disorder: A nonrandomized controlled pilot study. Journal of Affective Disorders, 195:172–179, 2016.

[30] Lukas Schilberg, Teresa Schuhmann, and Alexander T Sack. Interindividual variability and intraindividual reliability of intermittent theta burst stimulation-induced neuroplasticity mechanisms in the healthy brain. Journal of Cognitive Neuroscience, 29:1022– 1032, 2014.

[31] Maricor Soriano, Alessandra Araullo, and Caesar Saloma. Curve spreads-a biometric from front-view gait video. Pattern Recognition Letters, 25(14):1595–1602, 2004.

[32] Mitchell Adrien St. Pierre and Minoru Shinohara. Transcutaneous vagus nerve stimulation at nonspecific timings during training can compromise motor adaptation in healthy humans. Journal of Neurophysiology, 130(1):212–223, 2023.

[33] Yunqi Tang, Zhuorong Li, Huawei Tian, Jianwei Ding, and Bingxian Lin. Detecting toe-off events utilizing a vision-based method. Entropy, 21(4):329, 2019.

[34] Windsor KC Ting, Faïza AR Fadul, Shirley Fecteau, and Christian Ethier. Neurostimulation for stroke rehabilitation. Frontiers in Neuroscience, 15:1–11, 2021.

[35] Milka Trajkova, Nathaniel Green, and Minoru Shinohara. ‘StreamPoseML’ an end-to-end open-source web application and python toolkit for real-time video pose classification and machine learning. Journal of Open Source Software, 9(104):6392, 2024.

[36] Ole-Bjørn Tysnes and Anette Storstein. Epidemiology of parkinson’s disease. Journal of Neural Transmission, 124:901–905, 2017.

[37] Chunxiao Wu, Peihui Liu, Huaili Fu, Wentao Chen, Shaoyang Cui, Liming Lu, and Chunzhi Tang. Transcutaneous auricular vagus nerve stimulation in treating major depressive disorder: A systematic review and meta-analysis. Medicine, 97:1–8, 2018.

[38] Manal Zafar, Ariyana Bozzorg, and Madeleine E Hackney. Adapted tango improves aspects of participation in older adults versus individuals with parkinson’s disease. Disability and Rehabilitation, 39(22):2294–2301, 2017.

